# Genetic overlap between major depression, bipolar disorder and Alzheimer’s Disease

**DOI:** 10.1101/2021.05.01.21256220

**Authors:** Fernanda C. Dos Santos, Ana Paula Mendes-Silva, Yuliya S. Nikolova, Etienne L. Sibille, Breno Satler Diniz

## Abstract

**Background:** Mood disorders, including major depression (MD) and bipolar disorder (BD), are risk factors for Alzheimer’s disease (AD) and possibly share an overlapping genetic architecture. However, few studies have investigated the shared loci and potential pleiotropy among these disorders.

**Methods:** We carried out a systematic analytical pipeline using GWAS data and three complementary (genome-wide, single variant, and gene-level) statistical approaches to investigate the genetic overlap among MD, BD, and AD.

**Results:** GWAS summary statistics data from 679,973 individuals were analyzed herein (59,851 MD cases and 113,154 controls; 20,352 BD cases and 31,358 controls; and 71,880 AD cases and 383,378 controls). We identified a significant positive genetic correlation between MD and AD (r_G_ = 0.162; s.e. = 0.064; p = 0.012), and between BD and AD (r_G_ = 0.162; s.e. = 0.068; p = 0.018). We also identified two pleiotropic candidate genes for MD and AD *(TMEM106B* and *THSD7A)* and three forBD and AD (*MTSS2, VAC14*, and *FAF1)*, and reported candidate biological pathways associated with all three disorders.

**Discussion:** Our study identified genetic loci and mechanisms shared by mood disorders and AD. These findings could be relevant to better understand the higher risk for AD among individuals with mood disorders, and to propose new interventions.

## 1 INTRODUCTION

Mood disorders, including major depression (MD) and bipolar disorder (BD), are risk factors for Alzheimer’s disease (AD). History of MD or BD across the lifespan can double the risk of developing AD in older adults^1-4^. The risk is even higher in older adults with concurrent MD and mild cognitive impairment^5^ or those with persistent depressive symptoms^6^. Clinical and epidemiological evidence suggests that mood disorders and AD may share common biological abnormalities that, when triggered in individuals with mood disorders, lead to faster progression to AD^7^. Methodological challenges such as the need for large sample sizes with the longitudinal assessment of mood and cognition, over long follow-up time (ideally 10 years or more), and biospecimens collection limit the direct assessment of possible mechanisms related to the higher risk of AD among individuals with mood disorders^8,9^. An additional challenge is the high heterogeneity of MD, BD, and AD’s pathophysiologic mechanisms.

The availability of large datasets (e.g., Psychiatric Genomics Consortium (PGC), International Genomics of Alzheimer’s Project (IGAP), UK Biobank) with both clinical characterization and whole-genome analysis, together with advances in mathematical and analytical models, allow the investigation of the genetic overlap among complex disorders without demanding the assessment of both phenotypes in large longitudinal cohorts. Some of these methods, such as cross-trait linkage disequilibrium (LD) regression, bivariate causal mixture model, and pleiotropy-informed conjunctional false discovery rate^8,10^, employ mathematical and statistical models to summary statistics results of two different phenotypes to evaluate the genetic correlation and polygenic overlap while accounting for possible genetic pleiotropic effects.

Previous studies applying these statistical methods to investigate the genetic overlap between AD and mood disorders in the PGC and IGAP cohorts found no genetic correlation (LSDC measure of correlation effects) between AD and MD or between AD and BD^8^. Subsequent studies in non-overlapping samples (Generation Scotland’s Scottish Family Health Study (GS:SFHS) and UK Biobank cohorts) also failed to detect genetic overlap between AD and MD, either through genetic correlation or a complementary polygenic risk score (PRS) approach^11^. Nonetheless, Drange et al. (2019)^12^ recently found evidence of two pleiotropic loci for AD and BD using a pleiotropy-informed conjunctional false discovery rate method (conjFDR)^13^, despite not finding a significant genetic correlation between both conditions. Drange’s study highlights the importance of using complementary strategies to assess genetic overlap among psychiatric disorders, giving their genetic complexity and heterogeneity and the limitations of the currently available analytical methods^9^.

In the present study, we carried out a multi-level analytical approach to investigate the genetic overlap between mood disorders (MD and BD) and AD. We first applied a genome-wide statistical approach, followed by a single-variant and a gene-based network approach. Given that these phenotypes are primarily driven by regulatory networks composed of multiple genes, often with small effects^14^, we also evaluated the overlap of genes with minimal effects over each disorder and their shared biological pathways^15^. Since the individuals with mood disorders were on average younger than those with AD, we assumed that mood disorders would precede AD’s development in all analyses.

## 2 METHODS

### 2.1 Analyzed datasets

Summary statistics from three genome-wide association studies and meta-analyses were used to obtain candidate risk loci for mood disorders and AD. Summary statistics from MD and BD studies were extracted from the PGC2-MDD and PGC2-BD, respectively^16,17^. A summary statistics of an AD GWAS meta-analysis including the cohorts PGC-ALZ, IGAP and ADSP^18^ were assessed to account for AD candidate variants. There is few or no overlap of samples between mood disorders and AD databases. Supplementary material provides additional information about each dataset.

### 2.2 Genetic correlation

Linkage Disequilibrium Score Regression (LDSR) bivariate genetic correlations based on genome-wide SNPs (r_G_) were estimated for the pairs, AD vs. BD, and AD vs. MD using the LDSC software v1.0.0^8^. This method uses the relationship between GWAS test statistics (X^2^) and the linkage disequilibrium score of a given SNP to access the portion of heritability attributed to the same SNP in different studies and the genetic covariance of this given SNP across traits. The genetic correlation is calculated by normalizing the genetic covariance by the heritability estimates obtained from each trait (see supplementary material for additional information).

### 2.3 Polygenic overlap and pleiotropy

Polygenic overlap accounts for the fraction of genetic variants with a non-zero genetic effect that is causally associated with both traits over the overall number of variants identified as causal for each trait. Polygenic overlap was estimated using two methods. First, the polygenic overlap was estimated using a bivariate causal mixture model implemented in MiXer v1.2.0 software^19^. This method uses a bivariate causal mixture model, an expansion of the cross-trait LDSR method, in which a relaxed infinitesimal assumption is applied (see supplementary material for additional information).

We additionally used a pleiotropy-informed conditional false discovery rate (condFDR and conjFDR) to assess common variants associated with AD and mood disorders (pleioFDR v1.0.0 software)^13^. This method detects shared susceptibility loci in related phenotypes by applying a Bayesian method that tests the association of genetic variants to the principal phenotype when conditional on a second and related phenotype using the posterior probability of a false positive association^12,13^.

### 2.4 Gene-based GWAS and gene-based overlap

We performed a gene-based association approach to identify potential gene overlaps between mood disorder and AD using Multi-marker Analysis of GenoMic Annotation (MAGMA) v1.06 software^20^. Genes were annotated by considering SNPs’ position in a range interval containing 50 kb upstream and downstream the gene. We generated two gene lists for each disorder: A) a list of genes associated with the disorder under a 5% Bonferroni correction for multiple testing; and B) a broader list of all genes showing non-zero effect (i.e., nominal p-value < 0.05) (including those in List A) to guarantee that even genes with small effects would be evaluated. Lists A and B were compared between pairs of disorders, and a list of overlapping genes under each threshold was obtained. The gene model ‘multi’ was applied to obtain the association tests by the genes. Comparisons of overlapping among gene sets were performed considering two thresholds: 1) a Bonferroni corrected p-value (α=0.05), and 2) a non-zero effect (Z>|±1.96| or p<0.05). The lists of overlapping genes considering a non-zero effect for both disorders were used further as inputs for functional analyses.

### 2.5 Gene set enrichment analysis

Gene set enrichment analyses were performed on lists of overlapping genes among pairs of phenotypes (genes with non-zero effect) using FUMA v1.3.6^21,22^. In this analysis, all genes in Ensembl v92 were selected as background and a minimum of overlapping genes with gene sets ≥ 2 was used. Fifty-four tissue types from GTEx v8 were used to search for expression enrichment by tissue. Q-values were calculated using the Benjamini-Hochberg procedure for accounting for multiple testing.

### 2.6 Pathway Enrichment Analysis

To understand the biological processes affected by the overlapping genes between mood disorders and AD, we performed an enrichment map analysis to discover biological processes and pathways associated with the identified gene. For each given gene list, pathway and process enrichment analysis were carried out using g:Profiler (https://biit.cs.ut.ee/gprofiler) including the following ontology sources: GO Biological Processes, Reactome, and KEGG (Kyoto encyclopedia of genes and genomes). All genes in the genome were used as the enrichment background. Each network was visualized using Cytoscape 3.8.2 and the Enrichment Map (version 3.3)^23^. The Auto Annotate and Cluster Maker 2 apps were used for the analysis. The terms with a p-value < 0.05, q-value < 0.05, a minimum count of 3, Jaccard and overlap combined > 0.375 and edge cutoff ≥ 0.5 were collected and grouped into clusters based on their membership similarities. The p-values were calculated based on the accumulative hypergeometric distribution^24^ and q-values were calculated using the Benjamini-Hochberg method for multiple testing^25^. To further capture the relationship between the terms, the enriched terms were plotted in a network, where edges connected terms with a similarity > 0.5. Each node represents an enriched term and is colored first by its cluster ID generated using the Markov cluster algorithm (MCL)^26^ (see supplementary material for additional information).

## 3 RESULTS

### 3.1 Positive genetic correlation between mood disorders and AD

We initially selected three large recently published GWAS studies for MD^16^ (59,851 cases and 113,154 controls), BD^17^ (20,352 cases and 31,358 controls), and AD^18^ (71,880 cases and 383,378 controls) to obtain summary statistics and evaluate the genetic correlation between these conditions (Supplementary information). We found a small but significant genetic correlation between MD and AD (r_G_ = 0.162; s.e. = 0.064; p = 0.012), and between BD and AD (r_G_ = 0.162; s.e. = 0.068; p = 0.018). These results indicate that mood disorders and AD share causal variants, with small to moderate effect sizes.

### 3.2 Genetic overlap and pleiotropy between mood disorders and AD

We next investigated the extent of the genetic overlap between mood disorders and AD to capture the mixture of effect directions across shared variants. We estimated the number of shared variants and the correlation of effects (rho) only among them, using a bivariate causal mixture model (MiXer)^19^. MiXer models are optimized for variants with small effect sizes that do not reach GWAS significance^19^. AD shows a genetic architecture characterized by lower polygenicity than other complex disorders, with some variants, such as the APOE locus, showing very large effects^9,27^. Therefore, to better accommodate AD variants in the MiXer model, we removed variants mapping to APOE locus (chr19: 45,000,000-45,800,000 bp) for this analysis and fitted a model based only on SNPs that did not reach GWAS significance (see supplementary material for additional information)^19^. Our results indicated an estimate of 371 causal variants shared between MD and AD from a total of 27,636 causal variants (Table 1). Despite the low polygenic overlap, we found a very strong correlation of effects within shared causal variants (rho_12_ = 0.99) (Table 1).

**Table 1.**
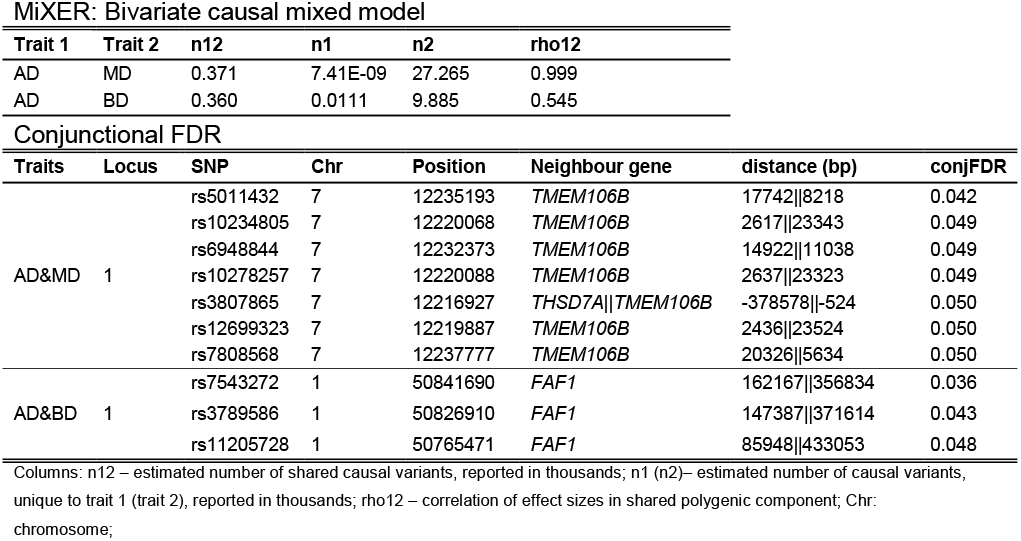
a) Results of MiXer cross-trait analysis for ‘Alzheimer’s disease (AD), Major depression disorder (MD) and Bipolar disorder for each thousand variants. b) Conjunction FDR; pleiotropic loci in AD and MD (AD&MD) and in AD and BD (AD&BD).

Given the MiXer model’s limitations, we further evaluated the genetic overlap by a pleiotropy-informed conditional false discovery rate (condFDR and conjFDR^13^). This method improves the detection of shared susceptibility loci in related phenotypes by applying a Bayesian method that tests the association of genetic variants to the principal phenotype when conditional on a second and related phenotype using the posterior probability false-positive association^13^. We identified one independent pleiotropic locus for AD and MD at a significance level of conjFDR<0.05 (Figure 1a and Table 1). This locus maps to chromosome 7 and contains 7 SNPs mapping nearby two genes, *TMEM106B* and *THSD7A*, which were mapped using SNPinfo Web Server^28^.

**Figure 1.**
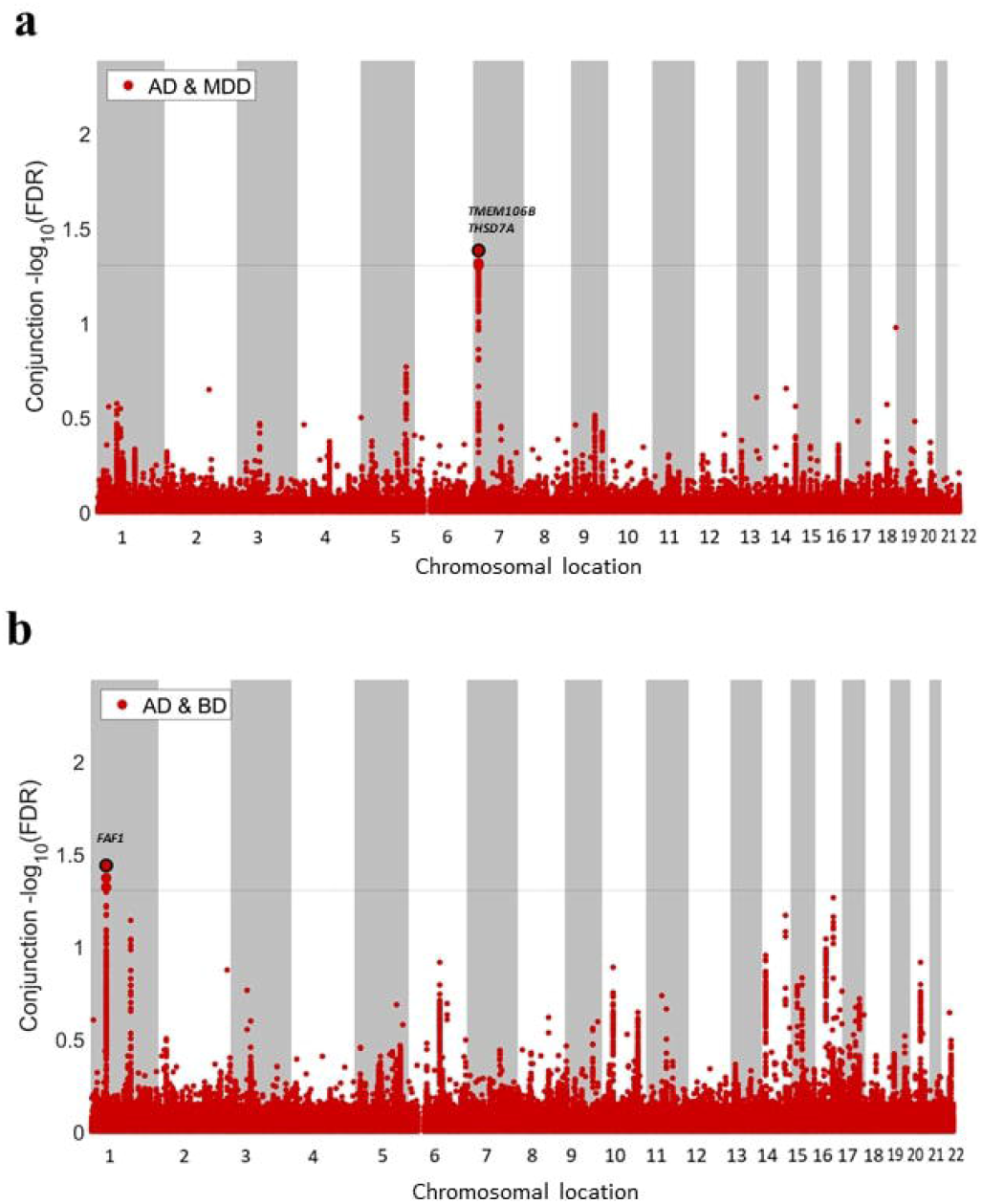
Conjunction conditional FDR Q value for SNP association with AD conditioned to a) MD and to b) BD. A genome-wide significant line (dashed) was drawn at –log10(0.05 FDR). Pleiotropic loci are circled in black.

For the BD and AD analyses, MiXer estimated 360 causal variants shared between BD and AD, from a total of 10,256 causal variants when considering both disorders (Table 1). A moderate positive correlation was found among shared variants (rho_12_=0.545). This result indicates that despite the low polygenic overlap between BD and AD, there is a significant but mixed correlation between the shared causal variants. The conjFDR also identified one pleotropic locus (conjFDR<0.05) mapping to the chromosome 1 and containing 3 associated SNPs, all mapping nearby the *FAF1* gene (Figure 1b and Table 1). Overall, these findings suggest a mixed pattern of effects among overlapping variants exists between BD and AD, in contrast with the MD and AD results. A full list of the loci found in this analysis can be found on Supplementary Table 1.

### 3.3 Gene overlaps between mood disorders and AD and related biological pathways

MAGMA analysis showed 220 and 35 genes associated with AD and MD, respectively (Supplementary tables 2 and 3). There were no overlapping genes between MD and AD after Bonferroni correction. However, considering all genes with non-zero effects over both disorders, we identified 158 genes overlapping genes (Supplementary table 4).

We included the 158 overlapping genes with non-zero effect found in MAGMA analysis to interrogate the affected biological pathways in the enrichment analysis. The Enrichment Map analysis^23^ returned 291 gene clusters for terms related to the cellular response to amyloid-beta, cell growth and differentiation, respiratory electron transport chain, synapse organization, immune-inflammatory response, membrane trafficking, gene and protein regulation, among others (Figure 2; the full result is available in Supplementary table 5). A highly interconnected pattern can be observed in this network, suggesting different sets of genes (with different effect sizes over MD and AD) impact biological pathways involved in both disorders, indicating potential mechanistic links between these disorders (Figure 2).

**Figure 2.**
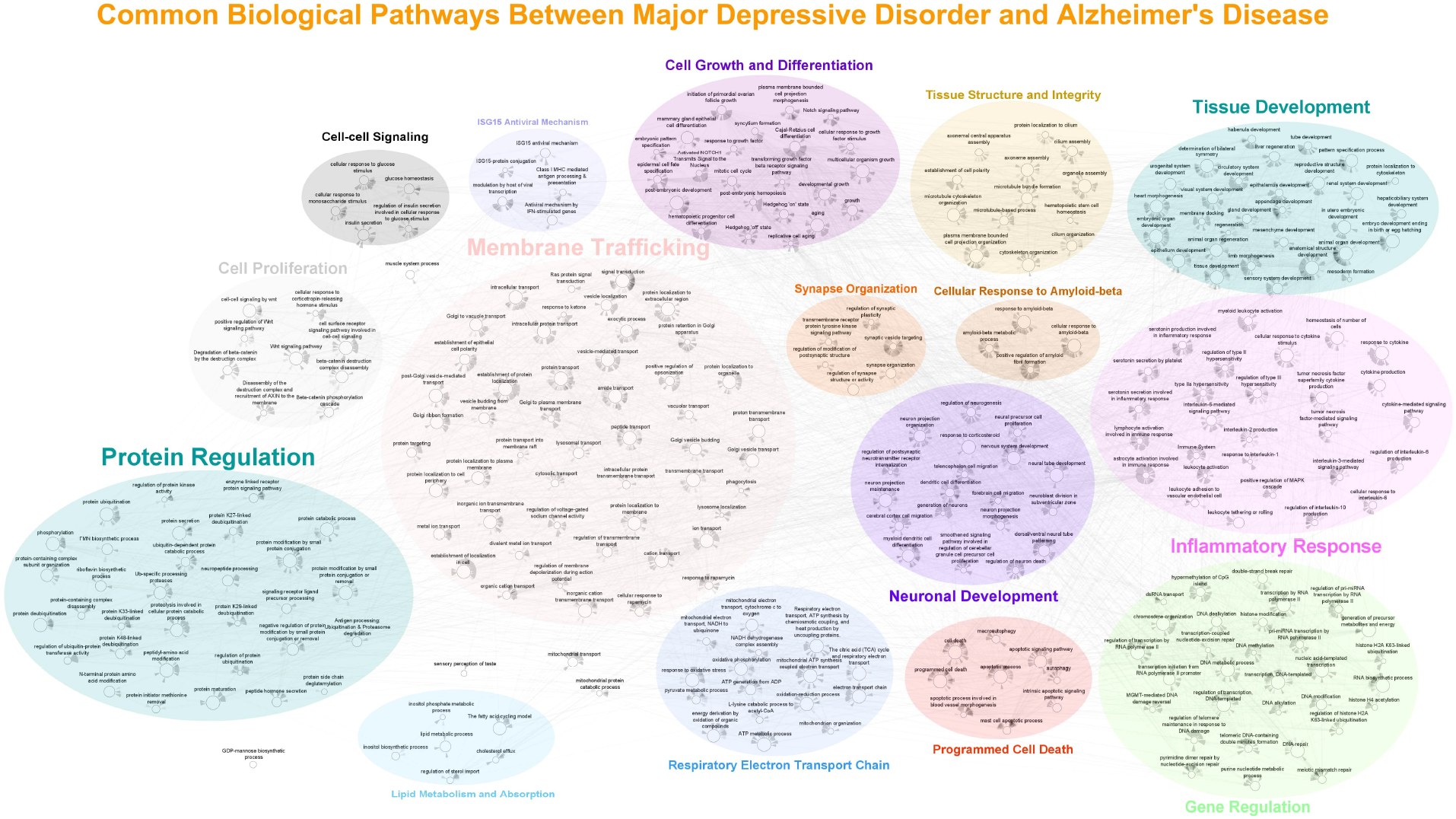
Common biological pathways between Major Depressive Disorder and Alzheimer’s Disease. A very interconnected pattern can be observed in this network, suggesting that sets of genes including genes shared between these disorders (with different effect sizes over MD and AD) impact a same biological pathway or even different biological pathways involved in these disorders. These genes could then indirectly influence the clinical overlap of them.

MAGMA analysis identified 245 and 220 genes associated with BD and AD, respectively (Bonferroni corrected p-value < 0.05) (Supplementary tables 2 and 6). We found 2 overlapping genes mapped to the same locus of chromosome 16: *MTSS2* and *VAC14*. Enrichment Map analysis detected 326 gene clusters related to cell cycle, cell growth and differentiation, cell-cell signaling, synapse organization, neuron death, respiratory electron transport chain, immune inflammatory response, membrane trafficking, gene and protein regulation, lipid metabolism and absorption, among others (p-value<0.05 and q-value<0.05) (Figure 3; the full result is available in Supplementary table 7). The six most enriched clusters involved inflammatory response, programmed cell death, protein and gene regulation, membrane trafficking, and tissue development (Supplementary table 7). The biological pathways analysis from overlapping genes between BD and AD showed a strong relationship between these clusters and neuronal development, neuron death, and synapse organization clusters. Other gene clusters showed a cellular senescence mechanism involved in cell cycle regulation and a programmed cell death stress-activated. As with MD and AD, a highly interconnected network can be observed when analyzing the overlapping gene sets between BD and AD (Figure 3).

**Figure 3.**
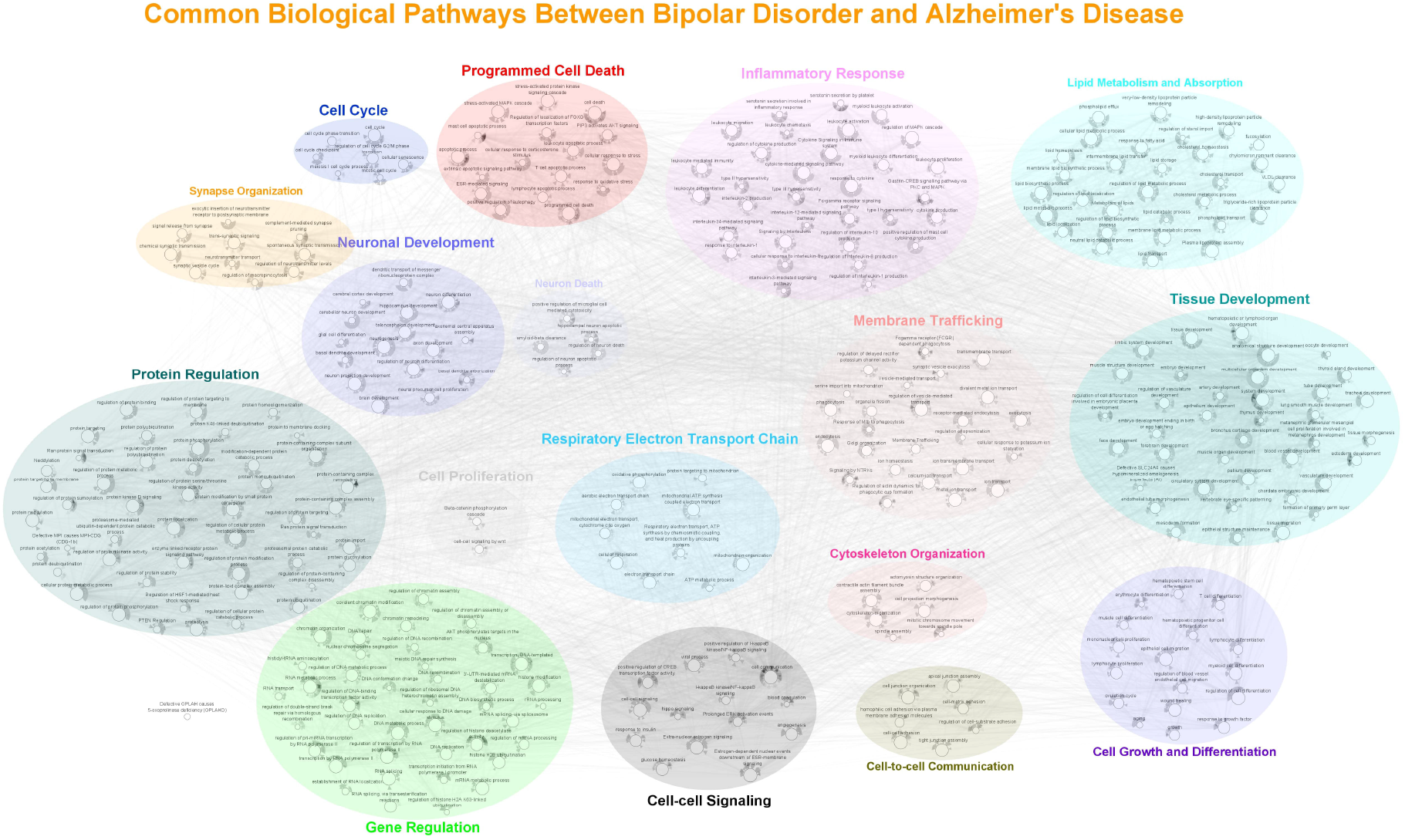
Common biological pathways between Bipolar Disorder and Alzheimer’s Disease. A very interconnected pattern can be observed in this network, suggesting that sets of genes including genes shared but with different effect sizes over BD and AD impact a same biological pathway or even different biological pathways involved in these disorders. These genes could then indirectly influence the clinical overlap of them.

### 3.4 Analysis of tissue expression of overlapping genes in mood disorders and AD showed enrichment for brain tissues

We investigated the tissue expression of overlapping genes between mood disorders and AD using the GTEX (v8) data in the FUMA GENE2FUNC application, covering 54 human tissues^21,22^. From the 158 genes overlapping between MD and AD, 156 were found in the GTEX database. These genes were mainly expressed in all nervous tissues except for the cerebellum, pituitary, and tibial nerve (corrected p-value < 0.05) (Figure 4a). Likewise, from the 279 overlapping genes between BD and AD, 278 were identified in the GTEX database. As with MD and AD analysis, these genes were mainly expressed in nervous tissues (Figure 4b). These results corroborate that genes related to mood disorders and AD are mostly involved in nervous system functions.

**Figure 4.**
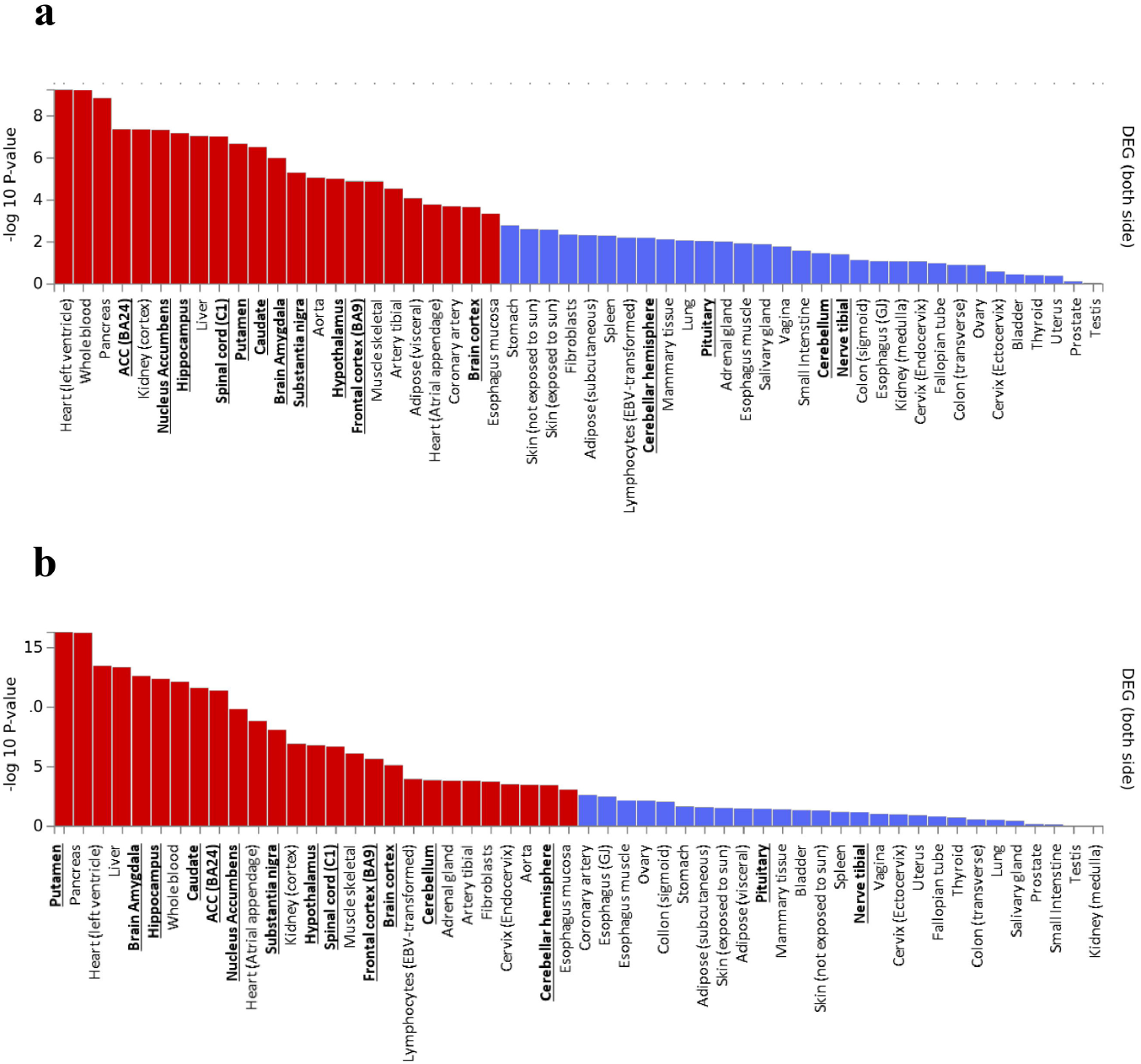
Pre-calculated differentially expressed genes (DEG) sets created using FUMA for a) MD and AD overlapping genes list and b) for BD and AD overlapping genes. Enrichment analyses were performed using GTEX v.8 54 tissue types database. DEG sets are obtained by a two-sided t-test per tissue versus all the remaining tissues. Significantly enriched DEG sets (P Bonferroni < 0.05) are highlighted in red. The - log10(P values) refers to the probability of the hypergeometric test. Neural tissues are highlighted in bold.

## 4 DISCUSSION

In this study, we applied a systematic analytical framework based on statistical methods and functional analyses to investigate the common genetic architecture between mood disorders and AD. Our main findings were a significant, positive genetic correlation between mood disorders and AD and the evidence of overlapping and pleiotropic loci among these disorders. To the best of our knowledge, our study is the first to provide a comprehensive analysis of potential overlapping genetic and biological pathways between mood disorders and AD and to identify pleiotropic loci contributing to MD and AD. Importantly, our analyses showed that the overlapping genes were mainly expressed in brain tissues.

In contrast with the previous studies^8,11,12^, we showed a significant genetic correlation among MD, BD, and AD. A possible explanation for our results was the use of larger publicly available databases compared to previous studies, increasing the power to detect statistically significant correlations of effects by cross-trait LDSR method. Using a single-variant approach, we found two candidate genes in a pleiotropy locus in MD and AD, expressed in the CNS and previously associated with essential functions described for both disorders. The Transmembrane Protein 106B (*TMEM106B*) is a protein-coding gene expressed in many tissues, including the brain. It is involved in dendrite morphogenesis, maintenance, and branching by regulating lysosomal trafficking and organization. Mutations in *TMEM106B* resulting in elevated expression of this protein have been identified as a risk factor for MD and AD in independent GWAS studies^16,29^. The second candidate identified was the Thrombospondin Type 1 Domain Containing 7A (*THSD7A*), a protein-coding gene involved in endothelial cell migration and cytoskeletal organization. It is highly expressed in the CNS, and variants of the *THSD7A* gene are associated with educational attainment^30^, depressive symptoms^31^, and AD^32^ in recent GWAS. We identified one pleiotropic gene for BD and AD using the same approach, the Fas Associated Factor 1 (*FAF1*). This gene codifies an ubiquitin-binding protein expressed in various tissues, including the nervous system. FAF1 initiates or enhances apoptosis through the interaction with the FAS antigen (TNFRSF6) and activation of caspase-8 and caspase-3^33,34^ and regulates necrosis after oxidative stress in neuronal degeneration^35^. *FAF1* has been associated with depressive symptoms and alcohol consumption in a GWAS^36^. These are important findings since they can lead to the development of interventions to mitigate the risk of AD among individuals with mood disorders.

Enrichment map analysis showed common biological pathways shared by MD and AD^23^. First, we identified biological processes involved in the cellular response to amyloid-beta. The abnormal processing and metabolism of the amyloid-beta protein and the secondary cellular responses to the amyloid-beta insult are viewed as early and primary pathophysiologic mechanisms of AD^37,38^. Other biological processes shared between MD and AD include regulating inflammatory response, neurotrophic support, neuronal death, and mitochondrial function. Abnormalities in these processes are described in AD and are well-established consequences of amyloid-beta pathology. Based on our results, we hypothesize the presence of a structured biological network linking MD and AD. In this scenario, individuals with MD are more susceptible to the abnormal metabolism and the downstream cellular responses to the amyloid-beta, leading to the activation of neurotoxic biological cascades, with negative consequences to brain health and higher risk of AD. Our results also partially explain why individuals with recurrent depressive episodes, those with severe and persistently high trajectories of depressive symptoms, and those with concurrent cognitive impairment display a highest risk of AD^39-41^.

There is less robust data on the association between BD and AD, but a recent meta-analysis shows that BD is a significant risk factor for AD, having a greater odds ratio than MD and AD^2^. Although the genetic correlation between BD and AD was similar to the one between MD and AD, our additional analyses showed a more significant genetic overlap between BD and AD than MD and AD, which can partially explain the higher risk of AD among individuals with BD. Enrichment Map analyses revealed three major pathway modules related to the genetic overlap between BD and AD. The first cluster is mostly related to the nervous system homeostasis maintenance and metabolic activity. Amid these pathways, amyloid-beta clearance, the calcium signaling pathway, serotonergic synapse, and neurotrophin signaling have been previously associated with AD physiopathology^42-44^. In a second cluster, the pathways were primarily related to immune response, cell growth and survival, and neuroendocrine pathways.

Interestingly, these pathway modules were highly interconnected and acted as an immune-endocrine-neuronal regulatory network. Also, several pathways related to intestinal absorption were extensively connected with inflammatory response and neuronal development clusters. These results suggest the involvement of a gut– immune-brain axis as a shared mechanism between BD and AD^45-50^. The third cluster of pathways involved cell communication, cell cycle regulation, apoptosis, and lipid metabolism, previously associated with both BD and AD pathogenesis^33,34,36,51-54^.

The current results should be viewed considering the study limitations. Although the use of summary statistics of GWAS is a convenient and useful method to investigate polygenic overlap, it does not provide the data for each individual in the original studies. Therefore, decisions taken on the original studies regarding study design and statistical methods could impact the current results. Second, the original studies’ sample characteristics may have influenced our results, e.g., the high heterogeneity of the original studies samples (composed of many cohorts), sex and age distribution, ascertainment of major depression, bipolar disorder, and AD caseness. The MiXer analysis results should be viewed with caution, given the genetic architecture of AD and its impact on the model fitting used to evaluate individual variants’ effects in AD. Finally, the choice to work with a list of genes with non-zero effect for the functional analyses (p-value uncorrected for multiple testing) could lead to the selection of genes with spurious associations to each disorder. However, to deal with this possible limitation, posterior steps including multiple testing corrections would mitigate the impact of this methodological choice and increase the confidence about the pathways and biological mechanisms found in the present study. A similar approach has been successfully adopted by Cui et al. (2018)^15^.

In summary, the multi-level approach to identifying the genetic overlap between mood disorders and AD revealed a complex network of genetic overlap affecting multiple biological pathways essential for the neuronal function and CNS homeostasis and hormesis (the adaptive responses of biological systems to challenges throughout the system). Our study identified possible mechanisms linking mood disorders and AD that can be relevant to the development of novel therapeutic interventions to mitigate the risk of AD among vulnerable individuals.

## Data Availability

Summary statistics used in the present study were publicly available online in the following links: Alzheimer's disease: Jansen et al. (2019) - https://ctg.cncr.nl/software/summary_statistics; Bipolar disorder: Stahl et al., (2019) - https://www.med.unc.edu/pgc/download-results/; and Major depression: Wray et al., (2018) - https://www.med.unc.edu/pgc/download-results/.

## ACKNOWLEDGMENTS

We thank the authors of Jansen et al., 2019^18^, Psychiatric Genetic Consortium Bipolar Disorder Working Group (PGC-BIP)^17^ and Psychiatric Genetic Consortium Major Depressive Disorder Working Group (PGC-MDD)^16,29^ for making summary statistics available. This work was supported by ‘CAMH’s internal funding mechanisms. Dr. Diniz is supported by NIH grants R01MH115953 and R01MH118311.

## AUTHOR CONTRIBUTIONS

F.C.D.S. contributed to study design and conducted all the genetic, computational and statistical analyses for genetic overlap, interpretation of results and wrote the manuscript draft. A.P.M.S. performed the enrichment analyses, interpretation and substantially contributed to the manuscript draft by describing enrichment analyses methods and findings. Y.S.N. and E.S. contributed to manuscript editing and critical intellectual input about the analysis and results. B.S.D developed the study concept, oversaw all analyses and all stages of manuscript preparation. All authors reviewed and approved the manuscript before submission.

## SUPPLEMENTARY FIGURE LEGENDS

**Supplementary Figure 1. Methodological flowchart**.

## SUPPLEMENTARY TABLE TITLES

**Supplementary table 1. Conditional and Conjunctional FDR for Alzheimer’s disease (AD) loci given Major depression (MD) (AD**|**MD); and for Alzheimer’s disease (AD) loci given Bipolar disorder (BD) (AD**|**BD)**.

**Supplementary table 2. Gene-based GWAS results for Alzheimer’s disease**.

**Supplementary table 3. Gene-based GWAS results for Major Depression**.

**Supplementary table 4. Overlapping genes for AD&MD (Spreadsheet 1) and AD&BD (Spreadsheet 2)**.

**Supplementary table 5. Node details for ErichmentMap analysis of overlapping genes between Alzheimer’s disease and Major Depression**.

**Supplementary table 6. Gene-based GWAS results for Bipolar Disorder**.

**Supplementary table 7. Node details for ErichmentMap analysis of overlapping genes between Alzheimer’s disease and Bipolar Disorder**.

