## Supplementary material for "Genetic overlap between major depression, bipolar disorder and Alzheimer’s Disease": SuppInfo_DosSantos_etal_2021.docx

### SUPPLEMENTARY METHODS

#### Analyzed datasets

Summary statistics from three genome-wide association studies and meta-analyses were used to obtain candidate risk loci for mood disorders and AD. Regarding mood disorders, summary statistics from MDD and BD studies were extracted from the PGC2-MDD and PGC2-BD, respectively^1,2^. A summary statistics of an AD GWAS meta-analysis including the cohorts PGC-ALZ, IGAP and ADSP^3^ were assessed to account for AD candidate variants.

- Alzheimer's Disease

Jansen et al. (2019)^3^ performed a GWAS of clinically diagnosed AD and AD-by-proxy for a 455,258 individuals (71,880 cases and 383,378 controls) obtained from four independent consortiums. Phase 1 sample included individuals from Psychiatric Genomics Consortium (PGC-ALZ), IGAP and the Alzheimer's Disease Sequencing Project (ADSP), totaling 79,145 individuals (24,087 clinically diagnosed LOAD and 55,058 cognitively normal controls). In phase 2, 47,793 AD-by-proxy cases and 328,320 Control-by-proxy from UK Biobank were analyzed. A meta-analysis of phases 1 and 2 (phase 3) identified 29 loci associated with AD, 16 of which were previously identified in Lambert, et al. (2013)^4^, and 13 which are new. Only summary statistics of phase 3 were used herein, obtained from https://ctg.cncr.nl/software/summary_statistics.

- Bipolar Disorder

Stahl et al. (2019)^2^ performed a GWAS meta-analysis in 32 PGC cohorts, totaling 51,710 individuals (20,352 BD cases and 31,358 controls). A follow-up analysis using data for 9,412 BD cases and 137,760 controls from an independent cohort. A combined analysis of both the discovery and follow-up samples identified 30 loci associated with bipolar disorder, 20 of which are new. Only summary statistics of the discovery phase were used herein, obtained at https://www.med.unc.edu/pgc/download-results/.

- Major Depression

Wray et al. (2018)^1^ performed a GWAS meta-analysis including seven MDD and major depression cohorts, totaling 480,359 individuals (135,458 cases and 344,901 controls). 44 independent loci were associated with major depression diagnose, brain anatomical differences and MD clinical features. Complete summary statistics are publicly available only for six out of seven cohorts (excluding 23andMe cohort), totaling a sample of 173,005 individuals (59,851 cases and 113,154 controls). Summary statistics can be obtained at https://www.med.unc.edu/pgc/download-results/.

#### Genetic correlation

Linkage Disequilibrium Score Regression (LDSR) bivariate genetic correlations based on genome-wide SNPs (r_g_) were estimated for the pairs, AD vs. BD, and AD vs. MD using the LDSC software v1.0.0 ^5^. This method uses the relationship between GWAS test statistics (X^2^) and the linkage disequilibrium score of a given SNP to access the portion of heritability attributed to the same SNP in different studies and the genetic covariance of this given SNP across traits. In a second step, genetic correlation is calculated by normalizing the genetic covariance by the heritability estimates obtained from each trait. Therefore, the LDSC method estimates the correlation in effect sizes across traits. The sign of the correlation indicates if shared genetic effects have predominantly the same direction; however, for traits in which there is polygenic overlap, but distinct directions of effects are observed, cross-trait LDSR may indicate an absence of genetic correlation (r_G_~0). In this case, methods accounting for polygenic overlap independently of correlation of effect would be more suitable.

#### Polygenic overlap and pleiotropy

Polygenic overlap accounts for the fraction of genetic variants with a non-zero genetic effect that is causally associated with both traits over the overall number of variants identified as causal for each trait. Polygenic overlap was estimated using two methods. First, the polygenic overlap was also estimated using a bivariate causal mixture model implemented in MiXer v1.2.0 software^6^. This method uses a bivariate causal mixture model, an expansion of the cross-trait LDSR method, in which a relaxed infinitesimal assumption is applied. MiXer models the additive effect of a given variant in traits A and B by a mixture of four bivariate Gaussian components estimated from 1) variants that do not have any effect on either trait; 2) variants that affect only trait A; 3) variants that affect only trait B; and 4) variants that affect both traits. An estimate of correlation of effect sizes is obtained within the shared variants component (4), accounting for genetically correlated traits^6^. For mathematical convenience, the MiXer model applies Gaussian distribution to model the effects of variants over the traits^6^, considering that an infinitely large number of variants each contribute a little to the characteristic (Fisher's infinitesimal model), which implies that a large fraction of causal variants would have effect sizes close to zero. This is an approximation that accommodates better some disorders than others. AD shows a genetic architecture characterized by lower polygenicity when compared to other complex disorders, where some variants show very large effects, such as the variants in the APOE locus^7,8^. In this case, a model considering two causal components of small and large effects would be more suitable, however, this emcompass a mathemathical complexity that has not been implemented in any software. In these specific situations, MiXer developers suggest that only variants that did not reach GWAS significance are modeled, as MiXer model, still provides a good fit for them^6^. In order to better accommodate AD variants in the MiXer model, for this analysis, we removed variants mapping to APOE locus (chr19: 45,000,000- 45,800,000 bp) and fitted a model based only on SNPs that did not reach GWAS significance (right censoring; z1max 5.45 --z2max 5.45), as recommended by MiXer developers^6^.

Second, common variants associated with AD and mood disorders were also assessed using a pleiotropy-informed conditional false discovery rate (condFDR and conjFDR), implemented in the pleioFDR v1.0.0 software^9^. This method is used to improve the detection of shared susceptibility loci in related phenotypes by applying a Bayesian method that tests the association of genetic variants to the principal phenotype when conditional on a second and related phenotype using the posterior probability of a false positive association^9,10^.

#### Gene-based GWAS and gene-based overlap

Second, overlapping genes between studies were obtained using MAGMA v1.06 software^11^. Genes were annotated by considering SNPs' position in a range interval containing 50kb upstream and downstream the gene. The gene model 'multi' was applied to obtain the association tests by the genes. Comparisons of overlapping among gene sets were performed considering two thresholds: 1) a Bonferroni corrected p-value (α=0.05) and 2) a non-zero effect (Z>|$\pm$1.96| or p<0.05). The lists of overlapping genes considering a non-zero effect for both disorders were used further as inputs for functional analyses.

#### Gene set enrichment analysis

Gene set enrichment analyses were performed on lists of overlapping genes among pairs of phenotypes (genes with non-zero effect) using FUMA v1.3.6^12,13^. In this analysis, all genes in Ensembl v92 were selected as background and a minimum of overlapping genes with gene sets ≥ 2 was used. Fifty-four tissue types from GTEx v8 were used to search for expression enrichment by tissue. Q-values were calculated using the Benjamini-Hochberg procedure for accounting for multiple testing.

#### Pathway Enrichment Analysis and Visualization using g:Profiler and Enrichment Map

To understand the biological processes affected by the overlapping genes between mood disorders and AD, we performed an enrichment map analysis to discover biological processes and pathways associated with the identified gene. For each given gene list, pathway and process enrichment analysis were carried out using g:Profiler (https://biit.cs.ut.ee/gprofiler) that is an online toolset with the following ontology sources: GO Biological Processes, Reactome, and KEGG (Kyoto encyclopedia of genes and genomes). All genes in the genome were used as the enrichment background. Each network was visualized using Cytoscape 3.8.2^14^, and the Enrichment Map (version 3.3), Auto Annotate and Cluster Maker 2 apps were used for the analysis. The terms with a p-value < 0.05, q-value < 0.05, a minimum count of 3, Jaccard and overlap combined > 0.375 and edge cutoff ≥ 0.5 were collected and grouped into clusters based on their membership similarities. More specifically, p-values were calculated based on the accumulative hypergeometric distribution^15^, and q-values were calculated using the Benjamini-Hochberg procedure for accounting for multiple testing^16^. To further capture the relationship between the terms, the enriched terms were plotted in a network, where edges connected terms with a similarity > 0.5. Each node represents an enriched term and is colored first by its cluster ID generated using the Markov cluster algorithm (MCL)^17^. The MCL simulates random walks on the underlying interaction network by alternating two operations: expansion and inflation. First, loops are added to the input graph. By default, each node's loop weight is assigned as the maximum weight of all edges connected to the node. Then, this graph is translated into a stochastic "Markov" matrix. This matrix represents the transition probabilities between all pairs of nodes, and the probability of a random walk of length n between any two nodes can be calculated by raising this matrix to the exponent n – a process referred to as expansion. As higher length paths are more common between nodes in the same cluster than nodes within different clusters, the probabilities between nodes in the same complex will typically be higher in expanded matrices. MCL further exaggerates this effect by taking entry wise exponents of the expanded matrix and then rescaling each column so that it remains stochastic, a process called inflation. Clusters are identified by alternating expansion and inflation until the graph is partitioned into subsets so that there are no longer paths between these subsets. The most statistically significant term within a cluster is chosen to represent the cluster^14^.
