## Supplementary figures and images for "Genetic overlap between major depression, bipolar disorder and Alzheimer’s Disease"

### Supplementary_figure1_workflowchart.jpg

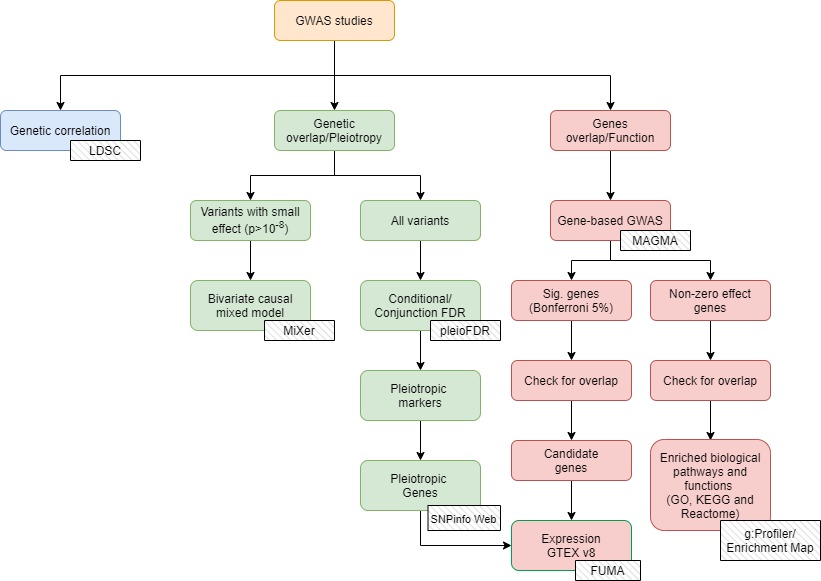
